# Validation of the GE Carescape V100 and Omron HEM-9210T by the AAMI/ESH/ISO 81060-2:2018 in multi-ethnic South-East Asian adult population

**DOI:** 10.1101/2022.11.23.22282694

**Authors:** Joel Chan, Cai Yue Sophia, Joshua Ng Yong Quan, Syuhaidah Binte Mohammed Noor, Shawn Wong Chong Yao, Chan Gerald, Christoph Chong Sheng, Wayne Ng Jia Wei, John C. Chambers, Marie Loh Chiew Shia, Theresia H. Mina

**Author notes:** **CORRESPONDING AUTHOR** Theresia H. Mina.

## Abstract

**Objective:** To ascertain the accuracy of 2 upper-arm blood pressure monitors, GE Carescape V100 Vital Signs and Omron HEM-9210T, used in a large, South-East Asian epidemiological cohort by the AAMI/ESH/ISO 81060-2:2018.

**Methods:** 149 participants were recruited from the ongoing Health for Life in Singapore (HELIOS) Study in multi-ethnic Singapore.

**Results:** 110 datasets were analysed. For criterion 1, the mean ± SD of differences for GE Carescape V100 was -3.5 ± 7.4/ -3.5 ± 7.4 mmHg (systolic/diastolic), and -4.2 ± 6.9/ -4.2 ± 6.9 mmHg (systolic/diastolic) for Omron HEM-9210T. For criterion 2, GE Carescape V100was -3.5 ± 5.6 / -3.5 ± 5.6 mmHg (systolic/diastolic), and -4.2 ± 4.5 / - 4.2 ± 4.6 mmHg (systolic/diastolic) for Omron HEM-9210T.

**Conclusion:** Both GE Carescape V100 Vital Signs and Omron HEM-9210T monitor passed the AAMI/ESH/ISO 81060-2:2018 validation standard.

## INTRODUCTION

The limited application of current human genetic studies due to the underrepresentation of Asians (1), and the urgency to identify etiological risk factors for complex diseases in Asia are our primary motivations to set the Health for Life in Singapore study (HELIOS), a prospective longitudinal population cohort comprising the multi-ethnic Asian adults living in shared environment in Singapore (primary ethnic groups: Chinese [76%], Malay [15%] and South Asians [9%]). Over the course of HELIOS Study protocol optimisation and expansion, we have used two upper-arm, automated sphygmomanometers, the GE Carescape V100 Vital Signs Monitor (referred as “GE”) and Omron HEM-9210T (referred as “Omron”). Although the Omron device has been previously validated against the 2013 ANSI/AAMI/ISO (2), no validation study has been conducted for the GE device. The main aim of this study was to validate both GE and Omron devices by the AAMI/ESH/ISO 81060-2:2018 standard (3), ensuring the accuracy and high-quality of the HELIOS research data.

## MATERIALS AND METHODS

### Study participants

HELIOS is approved by Nanyang Technological University Institutional Review Board (NTU IRB, reference: IRB-2016-11-030). This validation study was performed as one of the optional sub-studies offered to participants between August and December 2021. HELIOS participants were recruited from the general adult population of Singapore, with the inclusion criteria of being a Singaporean citizen or permanent resident aged 30-84 years, and the exclusion criteria of being pregnant or breast-feeding, having current acute illness or recent major surgery or mental incapacities preventing understanding of informed consent. There were 149 participants screened, and 110 participants completed the validation exercise. We excluded 7 participants due to arrhythmia and 32 due to blood pressure instability.

### Device

Both GE (GE Healthcare, Chicago, Illinois, US) and Omron devices (Omron Healthcare Co. Ltd., Kyoto, Japan) were equipped with a single cuff for arm circumference 23-33cm and 22-42cm, respectively. The reference manual, mercury-based sphygmomanometer is Yuwell (Jiangsu Yuyue Medical Equipment & Supply Co. Ltd.), with single standard cuff with 33cm upper limit, calibrated by the ISO/IEC 17025:2017-accredited ISOLAB Pte Ltd (Singapore).

### Procedure

The use of same arm sequential method was adopted (3). A dual-head stethoscope (3M, Saint Paul, Minnesota, US) was used to facilitate the auscultating and recording of the reference readings by 2 observers. We fulfilled the requirement for observers, supervisor and patient preparation as per the standard (3). As HELIOS is an adult cohort, we did not cover smaller cuff requirements for children and/or subjects aged 12-29 years. As we validated 2 devices concurrently and to ensure that there is no bias on the test device sequence, we randomly assigned participants to two arms (The first test device is Omron followed by GE, or vice versa).

### Statistical analysis

The data were analyzed as per the standard (3). Bland-Altman plots were created using SPSS version 26.0 (SPSS, Chicago, USA) according to the standard (3).

## RESULTS

In this validation study, the mean (SD) age of participants was 52.8 (11.9) years, and the male proportion was 43 (39.1%). There were 76 (69.1%), 10 (9.1%), 20 (18.2%) and 4 (3.6%) Chinese, Malay, South Asian and other ethnic Asian participants, respectively. The mean (SD) arm circumference was 25.6 (2.7) cm. **Table 1** describes the distribution of reference blood pressure measurements. **Table 2** summarizes the mean and SD from criterion 1 and 2 for the blood pressure difference between the test and reference device. The Bland-Altman plots do not illustrate any systematic bias **(Fig. 1)**.

**Table 1.**
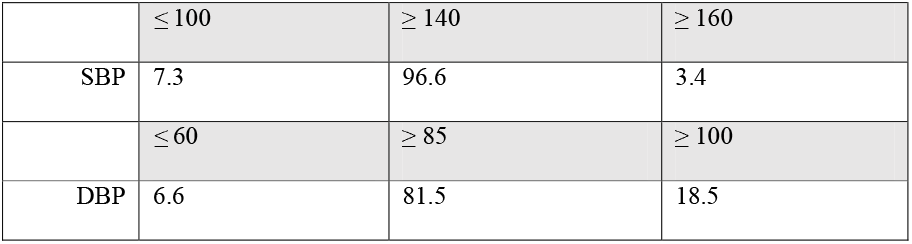
The distribution of reference blood pressure measurements (mmHg)

| | $\leq 100$ | $\geq 140$ | $\geq 160$ |
| --- | --- | --- | --- |
| SBP | 7.3 | 96.6 | 3.4 |
| | $\leq 60$ | $\geq 85$ | $\geq 100$ |
| DBP | 6.6 | 81.5 | 18.5 |

**Table 2.** Mean (SD) for the differences of the device and reference blood pressure values (mmHg). SBP= systolic blood pressure; DBP= diastolic blood pressure.

| Devices | Criterion | SBP | DBP |
| --- | --- | --- | --- |
| GE | 1 | $-3.5 \pm 7.4$ | $-3.5 \pm 7.4$ |
| | 2 | $-3.5 \pm 5.6$ | $-3.5 \pm 5.6$ |
| Omron | 1 | $-4.2 \pm 6.9$ | $-4.2 \pm 6.9$ |
| | 2 | $-4.2 \pm 4.5$ | $-4.2 \pm 4.6$ |

**Figure 1.**
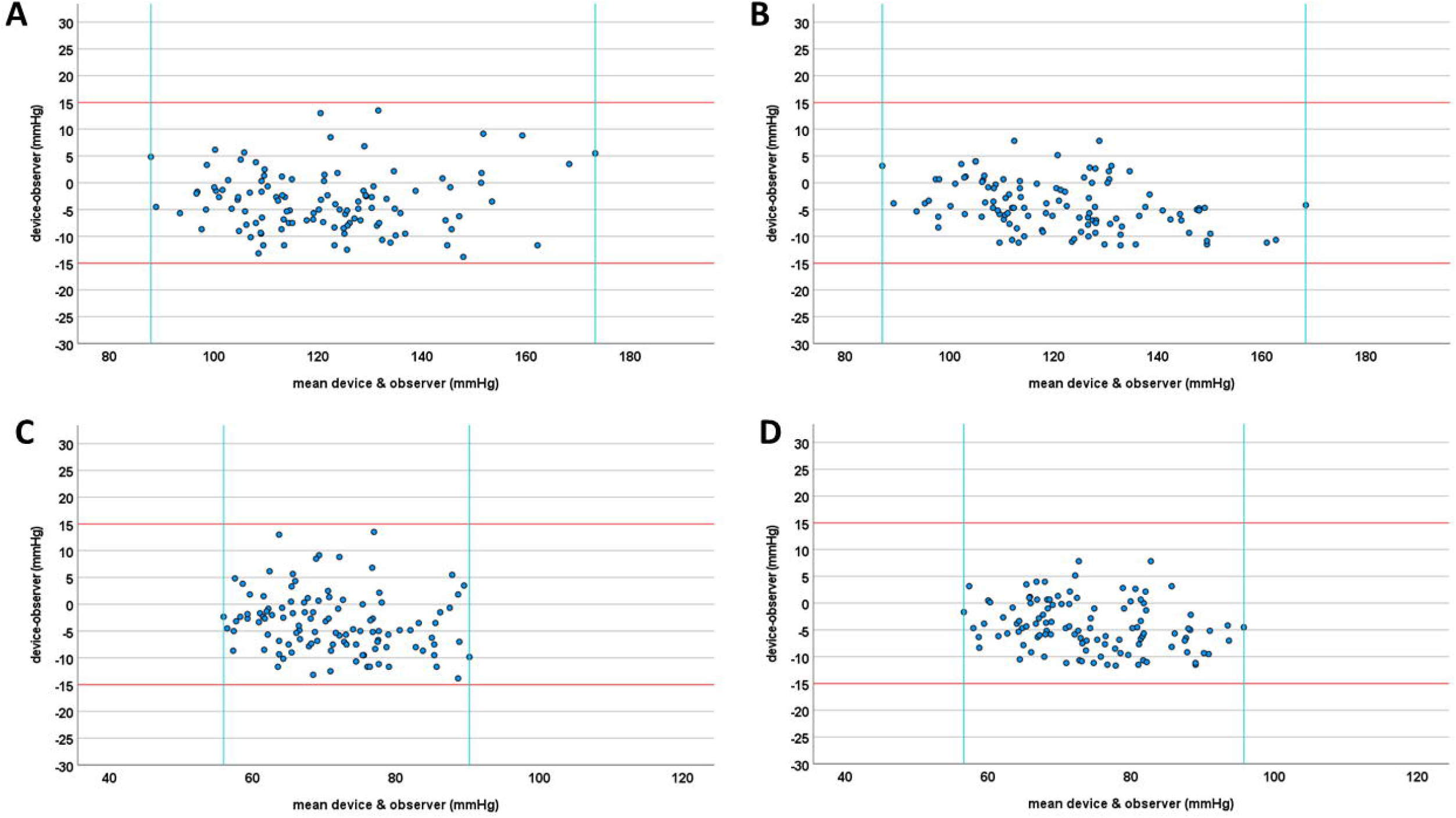
Bland-Altman plots of mean device and observer measurements of blood pressure values (X-axis) against the device-observer measurements (Y-axis). **A)** GE, systolic blood pressure (SBP); **B)** Omron, SBP; **C)** GE, diastolic blood pressure (DBP); **D)** Omron, DBP.

## DISCUSSION

This validation exercise demonstrated that both GE Carescape V100 Vital Signs and Omron HEM-9210T automated sphygmomanometer used in the HELIOS Study setting has passed the standard. As compared to the standard requiring 5% of reference systolic blood pressure measures ≥ 160 mmHg, the proportion observed in this validation exercise was slightly lower, although the diastolic blood pressure fulfilled the requirement. There could be 2 potential causes; first, most anti-hypertensive medications are more aggressive in lowering systolic than diastolic blood pressure (4,5), and second, although we did not observe bias towards healthier participants in the overall HELIOS Study, such bias could have been greater in this validation exercise as this exercise was offered as an optional sub-study.

In comparison with the earlier Omron validation study using 2013 standard (2), the mean (SD) difference in this study was also smaller, likely because we only focused on single-size mid-arm cuff and did not evaluate small or extra-large cuff. In the HELIOS setting, participants with very severe obesity (BMI ≤ 40 Kg/m^2^) and thus more likely to require extra-large cuff remains very rare (data not shown), but if critical numbers are reached as the study expands, future validation could repeat the exercise with extra-large cuff.

## CONCLUSION

Both GE Carescape V100 Vital Signs and Omron HEM-9210T automated sphygmomanometer passed the AAMI/ESH/ISO 81060-2:2018 standard.

## Data Availability

All data produced in the present study are available upon reasonable request to the authors

## ACKNOWLEDGEMENTS

We thank Dr. Terry Tong Yoke Yin, Ms. Shahidah Nur Shifa’ Binte Amran, Ms. Rebekah Wang, Ms. Choo Wee-Lin, and Mr. Lee Hong Kai for logistical and administrative support. We are grateful to the rest of the HELIOS study operational team for the overall support during data collection and the Lee Kong Chian School of Medicine Education Department for loaning the dual-head stethoscope.

## Notes

**CONFLICTS OF INTEREST** None declared.

**SOURCE OF FUNDING** JCC, MLCS and THM are supported by the intramural funding from NTU LKCMedicine, and the National Healthcare Group. JCC is supported by the Singapore Ministry of Health’s National Medical Research Council under its’ STaR funding scheme [NMRC/STaR/0028/2017]. THM received postdoctoral fellowship from the Singapore Ministry of Education – Research Scholarship Block and NTU LKCMedicine.

### Competing Interest Statement

The authors have declared no competing interest.

### Funding Statement

The is part of the Health for Life in Singapore (HELIOS) Study. HELIOS is supported by the Lee Kong Chian School of Medicine, Nanyang Technological University and National Healthcare Group, Singapore. J.C. is supported by the Singapore Ministry of Health National Medical Research Council under its STaR funding scheme [NMRC/STaR/0028/2017]. T.M. received postdoctoral fellowship from the Singapore Ministry of Education Research Scholarship Block and Lee Kong Chian School of Medicine.

### Author Declarations

Nanyang Technological University Institutional Review Board of Nanyang Technological University gave ethical approval for this work.

